# Integrative Genetics Analysis of Juvenile Idiopathic Arthritis Identifies Novel Loci

**DOI:** 10.1101/2020.09.01.20185603

**Authors:** Yun R. Li, Jin Li, Joseph T. Glessner, Jie Yang, Michael E. March, Charlly Kao, Jonathan P. Bradfield, Junyi Li, Frank D. Mentch, Huiqi Qu, Xiaohui Qi, Xiao Chang, Cuiping Hou, Debra J. Abrams, Haijun Qiu, Zhi Wei, John J. Connolly, Fengxiang Wang, James Snyder, Sophie Limou, Berit Flatø, Øystein Førr, Susan D. Thompson, Carl D Langefeld, David N Glass, Mara L. Becker, Elena Perez, Benedicte A. Lie, Marilynn Punaro, Debra K Shivers, Justine A. Ellis, Jane E. Munro, Carol Wise, Patrick M.A. Sleiman, Hakon Hakonarson

## Abstract

Juvenile Idiopathic Arthritis (JIA) is the most common type of arthritis among children, encompassing a highly heterogeneous group of immune-mediated joint disorders, being classified into seven subtypes based on clinical presentation.

To systematically understand the distinct and shared genetic underpinnings of JIA subtypes, we conducted a heterogeneity-sensitive GWAS encompassing a total of 1245 JIA cases classified into 7 subtypes and 9250 controls. In addition to the MHC locus, we uncovered 16 genome-wide significant loci, among which 15 were shared between at least two JIA subtypes, including 11 novel loci. Functional annotation indicates that candidate genes at these loci are expressed in diverse immune cell types. Further, based on the association results, the 7 JIA subtypes were classified into two groups, reflecting their autoimmune vs autoinflammatory nature.

Our results suggest a common genetic mechanism underlying these subtypes in spite of their different clinical disease phenotypes, and that there may be drug repositioning opportunities for rare JIA subtypes.

## Introduction

Juvenile Idiopathic Arthritis (JIA) is a chronic immune-mediated joint disease afflicting 1 in 10,000 North American children. Diagnosis of JIA is one of exclusion, since clinical features of JIA often overlap those of other rheumatologic diseases. The majority of autoimmune diseases (AID) are clearly-defined, for example type 1 diabetes (T1D) or Crohn’s disease (CD) which are clinically diagnosed based on results from serological or biopsy-proven results. However, JIA as well as other systemically manifesting diseases including rheumatoid arthritis (RA), systemic lupus erythematosus (SLE), and other collagen vascular diseases, represent a heterogeneous group of immune-mediated diseases that can be difficult to diagnose^1^. JIA causes severe joint pain and delays in therapy can result in short stature and joint disfigurement, prompting the need for new methods to identify early genetic or molecular diagnosis.

Although the sibling recurrence risk of JIA has been estimated to be between 15-30%^2,3^, few genetic risk factors have been clearly defined. Candidate and genome-wide association studies (GWAS) have identified genetic markers linked to JIA, including a strong association with the polymorphisms in the Class II MHC gene *HLA-DRB1*. While risk-associated HLA alleles, including *HLA-DR4*, are hypothesized to encode a shared disease epitope^4,5^, to date, the causative mutation(s) and disease mechanism are unknown. Because JIA is a relatively uncommon pediatric disease, few genome wide analyses have been performed compared to other autoimmune diseases; even less common are investigation across multiple JIA disease subtypes, of which there are seven, including Enthesitis-Related Arthritis (ENT), Polyarthritis RF Negative (PRFN), Polyarthritis RF Positive (PRFP), Oligoarthritis (OLG), Psoriatic Arthritis (PSO), Systemic Arthritis (SYS), and Undifferentiated Arthritis (UND).

Biological and molecular evidence based on the presence of serum autoantibodies and other molecular biomarkers of immunological defects suggest that the existing seven JIA subtypes are highly heterogenous. Some resemble predominantly seropositive autoimmune (AI) diseases, while others represent features of seronegative autoinflammatory (AIF) diseases. Whether genetic risk factors differ for AI Disease versus AIF-like JIA subtypes is unknown, especially since the traditional approach to genetic association studies is poorly powered to identify loci with subtype-specific effects in the presence of significant phenotypic heterogeneity.

The largest genetic association analysis in JIA, thus far, identified 14 susceptibility loci in oligoarthritis, the most common JIA subtype. Many of the loci reported are known AI disease signals, in part because the study used the immune-focused Immunochip genotyping platform (Illumina) that has been used to study over a dozen other immune-mediated diseases, including RA. A number of non-*HLA* single nucleotide polymorphisms (SNPs) are also associated with JIA, many of which are linked to other AI Diseases or AIF diseases AIFs^6-8^. This overlap is unlikely to be coincidental, as some JIA subtypes (polyarthritis and oligoarthritis) share serological and molecular features in common with other AI diseases such as RA, while findings in ENT and SYS represent more of the characteristics of AIFs and overlap with CD. Recent genomic studies and transcriptomic studies support this notion^9,10^.

Together, known loci explain less than a fifth of JIA heritability (13% of which are accounted for by signals across the HLA) ^8^. This contrasts with other polygenic AI Diseases like RA or T1D, for which >50% of heritability are explained by known loci. This “missing” heritability may be partly due to JIA’s phenotypic heterogeneity, as the clinical gold standard for JIA classification using the ILAR criteria currently splits patients into seven distinct subtypes based on sets of clinical features that are not distinct either from a mechanistic or therapeutic standpoint ^1,11^.

In addition, patients with JIA may require prompt pharmacologic intervention which is complicated by JIA often being a diagnosis of exclusion. Therefore, there is a clear impetus to identify genetic markers specific for JIA that can be used to separate JIA subtypes by disease mechanism. However, genetic analysis of a heterogeneous disease such as JIA requires a unique study design; for example, if a risk-associated SNP has subtype-dependent effects or if it has opposite effects on disease risk across subtypes, a “classic” case-control study treating all disease subtypes as cases of a single disease would lose power by introducing significant phenotypic heterogeneity. Given these differences in disease biology, it is plausible that genetic differences underlie AI Disease vs AIF JIA subtypes. Identifying markers that differentiate forms of JIA and potential biological mechanisms underscoring these associations will enable more prompt diagnosis and a classification system with greater clinical utility.

We hypothesized that the phenotypic heterogeneity in JIA is determined by genetic factors and sought to identify genetic associations that can more properly classify JIA patients. The definition of “genetic subtypes” has been successfully applied in other heterogeneous diseases, including breast cancer, for which genetically-targeted therapeutic options are now available^12-17^. In addition, recent work illustrates clear genetic differences between RF+ versus RF- RA that may play important roles in disease etiology ^18^, suggesting that by identifying molecular and genetically-defined RA subtypes helps shed light on disease mechanism. Finally, given that a large number of immune pathway modulators (e.g. TNF inhibitors; anti-IL1R antibodies) are already approved for use in other rheumatologic and immune-mediated disorders, including RA ^11,19-21^, knowing the genetic basis of JIA subtypes may allow for early and rapid introduction of already approved drugs to treat specific patient strata based on genetic definitions of molecular subtypes. Recent work focusing on JIA subtypes has started to uncover the genetic differences underlying these diseases with overlap in clinical presentations^9,22^. However, these studies have not systematically examined all seven subtypes of JIA in terms of genome-wide variants.

Here, we report a subset-sensitive GWAS study, including identification of 16 genome-wide significant loci which are specific to certain JIA subtypes or shared between specific subtypes. We conducted fine mapping of these loci to identify the potential causal variants and candidate genes at each locus. We further performed unsupervised clustering based on the association of these genetic loci with JIA subtypes. The integrative genetic analyses presented provide new insight into the biological differences between JIA subtypes.

## Results

### Identification of novel pleotropic JIA loci through hsGWAS

We compiled a JIA case-control cohort, that includes 1,385 JIA cases including all seven subtypes and 10,352 control subjects ascertained as having no history of any existing autoimmune or immune-mediated diseases. DNAs from all subjects were genotyped on two closely-matched Illumina Beadchip 550/610 platforms. The composition of the 7 subtypes within the JIA cohort is shown in **Fig. 1** and **Table S1**. A total of 507,092 SNPs in 1,245 cases and 9,250 controls passed quality control (QC) filtering.

**Figure 1.**
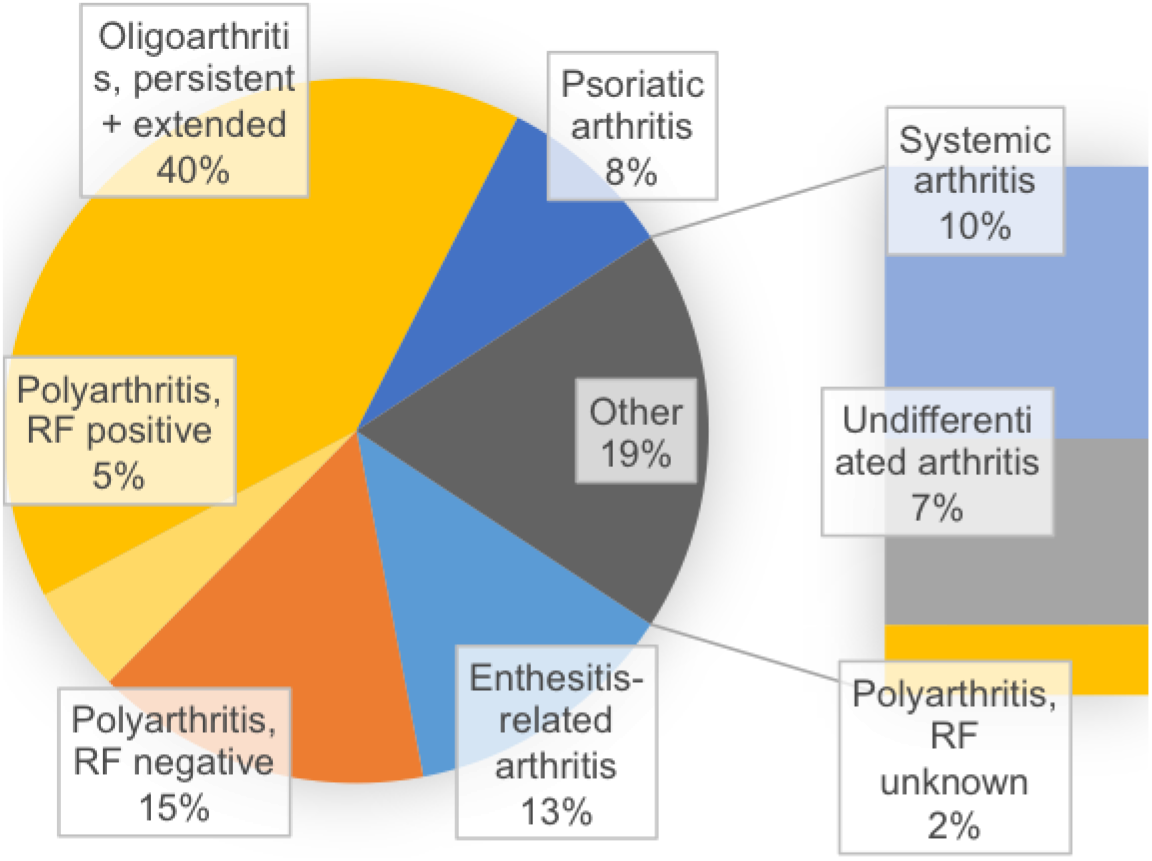
The composition of the subtypes in the JIA case-control cohort.

To optimize study power, we performed a heterogeneity-sensitive GWAS (*hsGWAS*)^23^ across the 1,245 JIA cases by splitting the case samples into the seven clinically-defined JIA subtypes and analyzed these subtypes and combinations thereof using the pool of shared non-AI disease samples. As described previously, the *hsGWAS* approach tests first every genotyped SNP passing QC to identify the most strongly associated disease combinations at each SNP, and then applies the DLM method for multiple-testing adjustment. We included the first 9 principal components (PCs) as covariates, which resulted in a genomic inflation factor (GIF) of 1.01 (**Fig. S1**), suggesting population stratification is well controlled. Our results showed that the majority of the association loci reported in previous publications were replicated in our study. Among the 121 association signals reported in GWAS catalog, including those at genome-wide marginal significance (P-value < 1e-05), 81.0% were replicated in our study at least at nominal significance (**Data file S1**).

We observed 16 loci surpassing genome-wide significance (GWS) (**Fig. 2**), including strong association signal at the MHC region (the 25–35 Mb region on chromosome 6 was considered as one association locus). Five out of the 16 genome-wide significant loci overlapped with previously reported autoimmune-disease loci (rs2066363, rs144844686, rs7660520, rs7731626, rs12203592) (**Table S2**), and the remaining 11 are novel GWS loci for JIA highlighted in **Table 1**. The regional association plots of these known loci and novel loci are shown in **Fig. S2** and **Fig. 3**, respectively. We further conducted conditional analyses conditioning on the index SNP at each locus to identify independent signals at each of the 11 novel loci, and found secondary independent association signal at three loci (3q28, 4q34.3, 7q11.22 and 18q21.1) (**Table S3**).

**Table 1.** The summary statistics of the independent loci reaching genome-wide significance.

| SNP | Chr | Pos | Candidate Gene | A1 | MAF | P_adj | Subtypes |
| --- | --- | --- | --- | --- | --- | --- | --- |
| rs2066363 | 1 | 82237577 | LPHN2 | T | 0.31 | 6.82E-11 | ENT,PRFN,PRFP,OLG,PSO,SYS,UND |
| rs144844686 | 2 | 234576970 | USP40 | T | 0.029 | 7.40E-09 | PRFN,OLG,PSO |
| **rs7636581** | **3** | **189781195** | **IL1RAP, CLDN1** | **A** | **0.12** | **6.82E-11** | **PRFN,OLG,PSO** |
| **rs13119493** | **4** | **180911259** | **intergenic** | **G** | **0.048** | **3.26E-08** | **PRFN,OLG** |
| rs7660520 | 4 | 183745321 | DCTD, TENM3 | A | 0.27 | 6.82E-11 | ENT,PRFN,PRFP,OLG,PSO,SYS,UND |
| rs7731626 | 5 | 55444683 | ANKRD55,IL6ST | A | 0.38 | 4.62E-10 | PRFN,OLG,PSO |
| rs12203592 | 6 | 396321 | IRF4 | T | 0.19 | 4.62E-09 | ENT,PRFN,OLG,SYS,UND |
| **rs114664970** | **6** | **40127169** | **LRFN2** | **C** | **0.012** | **1.03E-08** | **PSO,SYS** |
| **rs727845** | **7** | **67607209** | **intergenic** | **G** | **0.19** | **4.49E-08** | **ENT,PRFN,PRFP,OLG,UND** |
| **rs7042370** | **9** | **12785073** | **TYRP1** | **C** | **0.44** | **1.39E-09** | **ENT,PRFP,OLG,PSO,SYS** |
| **rs117572873** | **10** | **91997663** | **KIF20B** | **G** | **0.011** | **1.52E-08** | **PRFP,OLG** |
| **rs12795402** | **11** | **32255936** | **RCN1** | **C** | **0.34** | **4.19E-08** | **PRFN,PRFP,OLG,** |
| **rs17703382** | **11** | **30715593** | **MPPED2** | **G** | **0.014** | **2.76E-09** | **PRFP** |
| **rs147585949** | **16** | **20726695** | **ACSM1** | **A** | **0.014** | **1.98E-09** | **ENT,PRFN,PSO** |
| **rs11663074** | **18** | **45023793** | **SMAD2** | **C** | **0.17** | **6.82E-11** | **ENT,PRFN,PRFP,OLG,PSO,SYS,UND** |
| **rs138816451** | **22** | **43649657** | **SCUBE1** | **G** | **0.016** | **1.63E-09** | **PRFN,OLG,PSO, UND** |

CHR=chromosome; POS=Position (bp); A1=alternative allele; MAF=minor allele frequency; P\_adj=adjusted P-value; subtypes=associated subtypes

**Figure 2.**
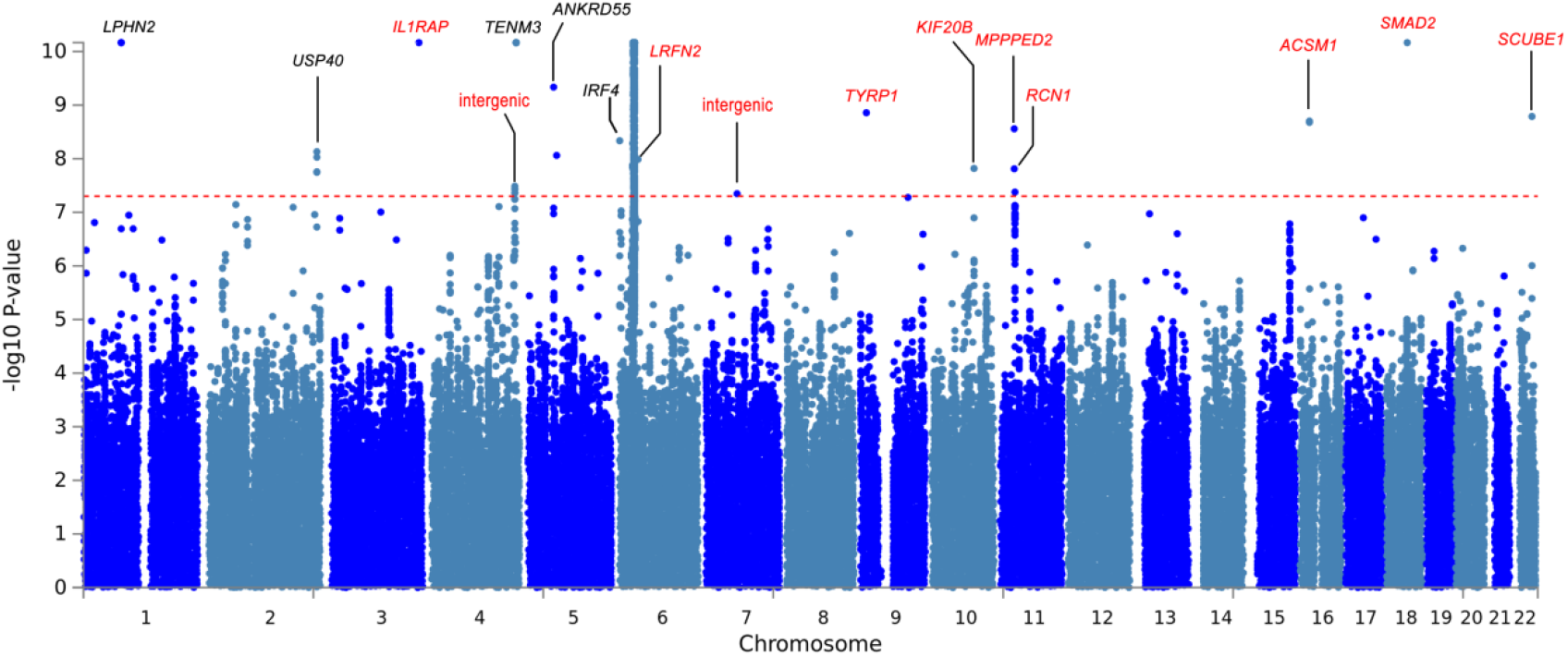
Association statistics in heterogeneity sensitive GWAS of JIA subtypes after multiple testing adjustment. The genome-wide significant loci are annotated with the candidate gene symbol and the novel loci are in red.

**Figure 3.**
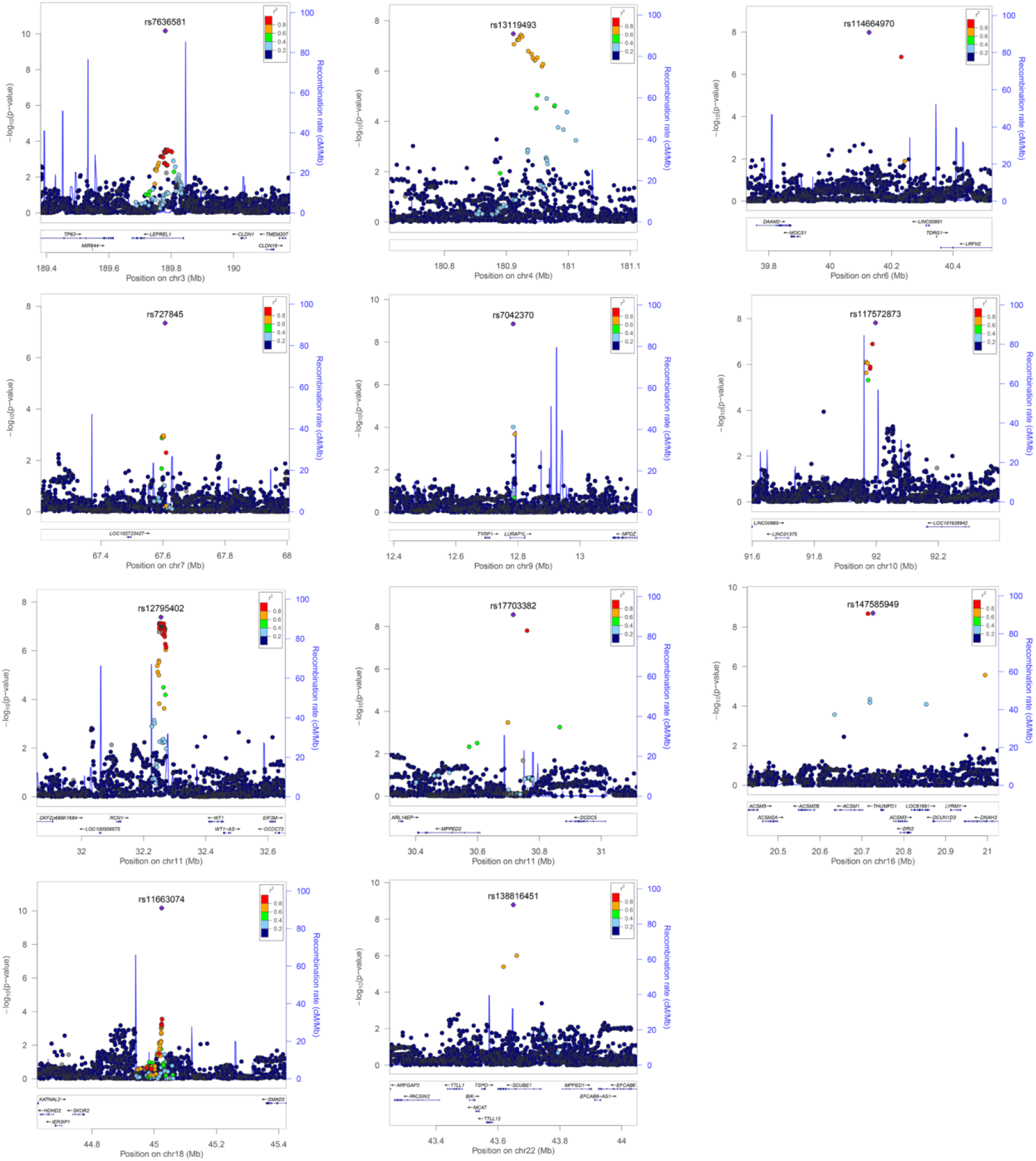
The regional association plots of novel JIA GWAS loci.

Among these 16 loci, only SNP rs17703382 is associated with only one JIA subtype, and all the others were shared among at least two JIA subtypes at GWS level (**Fig. S3**). In addition, we also observed 28 SNPs with genome-wide marginally significance (GMS) in our hsGWAS; and only 4 of them are associated with only one particular JIA subtype. The other 24 loci showed nominally significant associations with two or more JIA subtypes (**Table S4**).

### Replication of the novel JIA loci

Examining UK-biobank (UKBB) datasets, we found SNP rs7731626 is associated with rheumatoid arthritis at GWS. Interestingly, SNP rs12203592 is strongly associated with Rheumatoid factor > 16 IU/mL, while in our study, it showed a stronger association with PRFN and OLG than the other JIA subtypes. In addition, SNP rs7660520 showed association with Psoriasis at GWS and with Psoriasis arthropathy. In our dataset, this SNP showed associations with multiple JIA subtypes including PSO. A novel GWS SNP rs114664970 is associated with Chronic sinusitis in UKBB, the severe cases of which could be symptoms of autoimmune diseases (**Table S5**). We conducted another *in silico* replication by checking the association of these GWS loci with JIA in the immunochip data which was reported by Hinks et al. ^8^ At eleven out of 16 loci, the P-value of the top associated SNP is of nominal significance. Only modest replication was achieved, likely due to sparse SNP coverage at those loci which are not well-known immune loci (**Table S6**).

### Fine-mapping and functional annotation of the novel loci

For each of the novel loci, we conducted fine-mapping using the software, FINEMAP. By applying a threshold of posterior probability of 0.2 and original disease association P-value <5e-05, 11 SNPs were identified as potential causal variants, including index SNPs at 8 loci and 3 non-index SNPs (**Table S7**). We conducted functional annotation of all the index SNPs, the potential causal variants and the lead SNPs at GMS loci in ENCODE and ROADMAP epigenomics databases, and found that many of them overlap with chromatin marks or DNAse hypersensitive sites, likely playing a role in regulating target gene expression (**Fig. S4**).

We mapped these index SNPs to candidate genes based on eQTL and Hi-C data (**Fig S5, S6 and S7**), and the most-likely candidate genes at the GWS loci are indicated in the Manhattan plot (**Fig. 2**). Strong eQTL relationship was observed between SNP rs7731626 and gene ANKRD55 in different immune tissues and immune cell types. Fourteen candidate genes were expressed in immune cell types at medium to high level based on data extracted from the DICE database, including naïve B cells, naïve CD4 T cells, naïve CD8 T cells, Monocytes and NK cells (**Fig. S8**), suggesting both innate immunity and adaptive immunity are involved in the pathogenesis of JIA.

### Association of HLA alleles with JIA subtypes

Based on SNP genotype at the MHC region, we further imputed classical HLA-A, HLA-C, HLA-B, HLA-DRB1, HLA-DQB1, HLA-DQA and HLA-DPB1 alleles and examined their association with each JIA subtype (**Table S8**). We found highly significant association between HLA_B_27 and ENT as expected. HLA_DQB1_04 and HLA_DRB1_08 showed consistent significant associations across JIA subtypes of ENT, PRFN, OLG, PSO and UND. Additional HLA alleles were associated with JIA subtypes surpassing multiple-testing adjusted significance threshold. Further studies of the HLA class I and class II loci based on direct sequencing of the MHC region are underway to confirm these associations and to examine the HLA associations observed in more depth.

### Molecular clustering of JIA subtypes based on GWS loci

Previous biomarker studies suggested the autoimmune versus autoinflammatory nature of JIA subtypes. To investigate the relationship between JIA subtypes, we conducted unsupervised clustering based on the significance P-value of the GWS SNPs and GMS SNPs in hsGWAS. OLG and PRFN are clustered together as expected, with PSO being an addition into this cluster; the second cluster comprises SYS, ENT and UND; PRFP is the third cluster. We further incorporated the magnitude and direction of SNPs effects (reflected by association coefficient beta) into the clustering using tSNE algorithm. Using this approach, the 7 JIA subtypes are divided into two large clusters. SYS, ENT, UND and PRFP fall into a single cluster. The other cluster is composed of OLG, PRFN and PSO (**Fig. 4**), which is consistent with the notion that OLG and PRFN represent the classical AID but SYS is leaning toward the AIF diseases.

**Figure 4.**
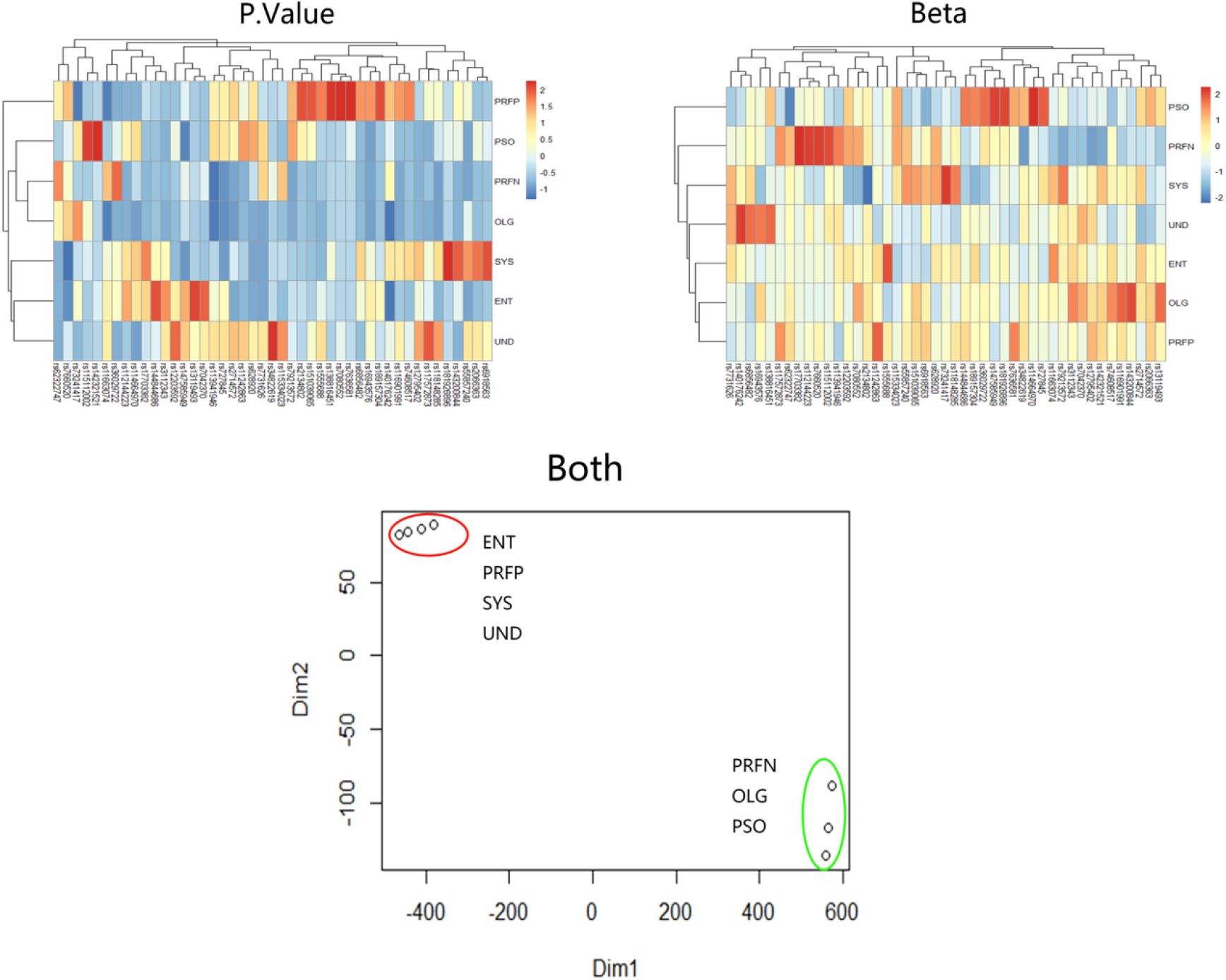
Clustering of JIA subtypes across the GWS and GWM loci on the basis of disease-specific association. The plots shown are agglomerative hierarchical clustering across 7 JIA subtypes on the basis of multiple-testing adjusted P-value from hsGWAS; agglomerative hierarchical clustering across 7 JIA subtypes on the basis of association beta from hsGWAS; and t-SNE clustering across 7 JIA subtypes on the basis of multiple-testing adjusted P-value and beta from hsGWAS.

### Pathway enrichment and network analysis of GWS loci

To better understand how these loci may contribute to JIA etiology, we further performed pathway enrichment analyses and protein-protein interaction (PPI) network analysis. We first looked into the most-likely candidate genes of the 16 GWS loci using the over-representation analysis, the KEGG pathway “Th17 cell differentiation” is significantly over-represented with four candidate genes “ SMAD2, IRF4, IL1RAP, IL6ST “ in this pathway (**Table S9**). The candidate genes also fall into several GO biological processes centered on immune functions, but these gene sets did not pass multiple testing correction (**Table S10**). Then investigating the whole hsGWAS results, we found the enrichment of 81 KEGG or BIOCARTA or REACTOME pathways (**Data file S2**) with pathways relating to autoimmune diseases ranking at the top. These top ranked pathways mostly were driven by the genes at the HLA locus on chromosome 6. PPI network analysis revealed the extensive interactions between immune genes centered on TNF, NOS1 and HLA genes (**Fig. S9**).

### Identification of drug targets with drug repurposing opportunities among the JIA associated loci

We searched several drug target gene databases rugBank (https://www.drugbank.ca/), Drugcentral (http://drugcentral.org/) and PharmGKB (https://www.pharmgkb.org/) and found candidate genes at multiple GWS loci are known targets of existing drugs which could be used for JIA treatment, including several that have been used for arthritis, such as Diflunisal, methotrexate, cyclosporine and diclofenac. (**Table S11**). The finding of arthritis drugs impactinggenes at GWS loci in our study supports the biological relevance of our study. In addition, it is interesting that the Hi-C data revealed evidence forchromatin interaction between rs7636581 and IL1RAP, suggesting that IL1RAP may be a candidate target gene of this locus. In regards to existing therapies, IL-1 antagonists (and IL-6 blockade) have already been in use for the SYS subtype and have been transformative in treating this sub-type of JIA^24^. In our analysis, the GWA signal is most significant for OLG and PSO, suggesting the potential application of IL-1 blockade in treatment of these subtypes of JIA in addition to SYS. Another interesting signal is the association of rs7731626 with IL6ST, encoding Glycoprotein 130 (GP130), a coreceptor for many other cytokine receptor complexes besides IL-6^25^. This locus is associated with multiple JIA subtypes. SYS subtype has been reported to be responsive to IL-6 blockade^26,27^. These observations suggest that therapies currently only focused on systemic forms of JIA for this target, may also be effective more broadly for other JIA subtypes. The association of rs138816451 with SCUBE1 on chr22 is intriguing as SCUBE1 has been implicated in playing a role in angiogenesis and is expressed and bound to the surface of endothelial cells^28^. There is a report suggesting that monitoring SCUBE1, SCUBE3 and VEGF levels in serum could be a biomarker for angiogenesis^29^, which is one of the pathogenic processes involved in psoriasis and arthritis, suggesting that drugs that block angiogenesis may be effective for treating arthritis and psoriasis/psoriatic arthritis^30-32^. The candidate gene SASH1 on chr6 is involved in pathological remodeling processes involving the endothelium and has been implicated in smoking induced atherosclerosis^33^ and endothelial response to TLR4 activation^34^. The rs151039065 association with MYCBP2 encodes an E3 ubiquitin ligase which has been shown to play a direct role in sphingosine-1-phosphate (S1P) signaling through mTOR and cAMP activation^35,36^. S1P receptor modulator/inhibitor drugs such as fingolimod and ozanimod have been approved for autoimmunity, including multiple sclerosis^37,38^.

## Discussion

JIA is an important chronic autoimmune disease among children, however, it has not been as well studied as many of the other autoimmune diseases, such as RA and T1D, mostly due to the limitations of sample size and its clinical heterogeneity. To address these limitations, we conducted a hsGWAS accounting for the phenotypic heterogeneity across multiple cohorts and identified novel pleiotropic loci shared among multiple JIA subtypes. We illustrated genetically how these loci may impose joint or disparate effect on the susceptibility of JIA disease subtypes.

The signaling pathway, “TH17 cell differentiation”, is significantly enriched among the candidate genes at the GWS loci, including IL1RAP, IL6ST, IRF4, and SMAD2. The rs11663074 association signal at 18q21.1 is a novel genome-wide significant locus of JIA. It is located 3.7kb from the 5’ of RP11-157P23.2, but the candidate gene is more likely to be SMAD2, which encodes a protein product belonging to the signal transducer and transcriptional modulator SMAD family. The protein complex of SMAD2/SMAD4 or SMAD2/SMAD4 is activated by TGF-beta (transforming growth factor) and by activating type 1 receptor kinases, and then functions as a transcriptional modulator, being involved in transcriptional regulation of the expression of TGF-beta target genes. TGF-beta signaling and SMAD2 play critical roles in Th1 cell development and in the generation of TH17 cells which can drive the development of autoimmune diseases^39-41^. The rs80142631 SNP at the SMAD2 locus has been reported to be associated with eosinophil counts in the population of European ancestry^42^. In our study, the minor allele C of SNP rs11663074 showed consistent protective effect across 7 subtypes. SNP rs7636581 is located in the intron of the LEPREL1 gene, however, the expression level of LEPREL1 is low in immune cell types and Hi-C data showed the chromatin interaction between rs7636581 and the IL1RAP gene region. Considering the known role of IL1RAP in immune cell types, it is more likely to be a candidate gene contributing to JIA pathogenesis. The protein product of IL1RAP belongs to the interleukin 1 receptor complex, involved in activation of IL1-responsive genes including genes in the NF-kappa-B signaling pathway. Both NF-kappa-B signaling pathway genes and other genes downstream of IL-1 play critical role in Th17 cell differentiation. Th17 cells are a lineage of CD4+ T cells, secreting cytokines IL-17A and IL-17F, which are involved in the pathogenesis of both autoimmune diseases (such as rheumatoid arthritis), and inflammatory diseases (such as inflammatory bowel disease). The other two candidate genes in the “TH17 cell differentiation” pathway are IL6ST and IRF4, both of which have been associated with autoimmune and autoinflammatory diseases in previous publications, including Crohn’s disease and RA.

Autoimmune and autoinflammatory diseases are two interrelated categories of systemic immune disorders that occur without exogenous pathogens. Both involve pathological processes in which the immune system is chronically activated and targeted against the patients’ own bodies, most notably including the musculoskeletal system and the skin, eventually resulting in tissue inflammation. A fundamental difference between these two types of diseases exist in the effectors mediating the immune action. In AIFs, the innate immune system directly causes acute inflammation; while in AI diseases, the innate immune system activates the adaptive immune system and then in turn leads to systemic inflammation. The existing seven JIA subtypes could be grouped, based on biological and molecular evidence, into two major classes—subtypes that represent either predominantly seropositive AI diseases or seronegative AIF diseases. Recent genomic studies have begun to reveal the genetic characteristics of JIA subtypes. GWAS and dense genotyping of autoimmune disease loci suggest that there is no significant overlap of genetic loci between SYS and other types of JIA mainly OLG and RFNP ^8,43^, and transcriptome analysis by RNA-seq further emphasized the similarity of SYS to autoinflammatory diseases, such as Crohn’s disease, while OLG and RFNP displayed biomarker-expression more characteristic of autoimmune diseases.^10^ Another JIA subtype RFPP both clinically and genetically resembles adult RA, especially early onset RA.^22^ Our clustering based on the strength and significance of association between each GWS locus and each JIA subtype is consistent with the above reports.

To manage the symptoms of JIA, various treatments have been developed; however, these therapies are of limited effectiveness as they are not based on the underlying causes of the disease but rather aimed at alleviating symptoms. In particular, therapies for the rare subtypes of JIA do not exist in part due to the limited understanding of the pathological basis of JIA and the ambiguity in JIA-subtype classification. IL-1 signaling plays important roles in regulation of pro-inflammatory reactions that are involved in various auto-inflammatory diseases^44,45^. IL-1 antagonists are available in the clinic, but they have not been explored much in JIA besides the SYS subtype. In adult psoriatic arthritis, IL-1 blockade has been shown to be effective^24^. In our JIA GWAS, rs7636581 is associated with IL1RAP with greatest contribuions from OLG and PSO subtypes, suggesting a rationale for looking at this locus more closely with respect to being a therapeutic target and informative biomarker of IL-1 signaling. Furthermore, IL1RAP acts as an accessory protein for not only IL-1 receptor but also for signaling through IL-33 and IL-36 (alpha, beta, and gamma) in relevant cell types^46^. Both IL-36 and IL-1 contribute to the pathobiology of psoriasis, suggesting that IL1RAP and its associated IL-1 and IL-36 signaling molecules may serve as biomarkers and therapeutic targets for JIA. There are therapeutic antibodies being pursued for both these cytokine pathways, with promising data from a study showing that anti-IL-36 is effective in a severe form of psoriasis^47^. Similarly the association of IL6ST with multiple JIA subtypes suggests the potential repurposing of anti-IL6 for at least several JIA subtypes. Patients with SYS have shown significant improvements in the signs and symptoms following treatment with tocilizumab for IL-6 inhibition in clinical trials, and the efficacy and safety of IL-6 inhibition in treating SYS patients have also been confirmed^26,27,48^. Tocilizumab has been demonstrated to improve patients’ symptoms in a pivotal phase III trial on polyarticular JIA^49^. Sarilumab, which is another antibody to the IL-6R, has undergone testing for polyarticular JIA and sJIA^50,51^. Our hsGWAS results suggest additional subtypes of JIA patients could potentially benefit from the treatment of IL-1 and/or IL-6 blockade. The novel GWS association of SCUBE1 with several JIA subtypes implicates the critical role of angiogenesis in JIA pathogenesis^30,52^. Some anti-inflammatory drugs, such as anti-TNF, anti-IL-6, have dual roles in blocking both inflammation and angiogenesis^53,54^. Drugs that target angiogenesis/vascularization (e.g. anti-VEGF, anti-TIE2 and anti-angiopoietins) may also have a role for these autoimmune diseases^55^. Indeed, case reports indicate that patients who were treated with anti-VEGF for cancer also showed dissolution of their psoriasis/psoriatic arthritis^56^. Similarly, drugs that target the mTOR pathway (i.e. the rapalogs and PI-3 kinase inhibitors)^57^ and drugs that modulate cAMP via phosphodiesterases (mainly PDE4 inhibitors such as aprelimast, crisaborole and roflumilast) are also drugs approved for inflammatory/autoimmune conditions^58^, with our genetic association data suggesting they may have a role in JIA.

One limitation of our study is that the sample size is not big in comparison with other GWAS of complex human diseases. However, pediatric disease expressing phenotypes in early life are typically with much stronger genetic signal than diseases presenting later in life that are often critically impacted by gene-environment interactions. Assuming a log-additive (multiplicative) regression model and a population disease frequency of 0.01%, our study design is well-powered (> 0.8) to detect a common SNP (MAF > 0.25) with an odds ratio > 1.15 and modestly-powered (> 0.5) to detect a SNP with MAF>0.05 and odds ratio (OR) > 1.20. Given that average OR of GWAS SNPs identified previously in oligoarthritis (the most common JIA subtype) is between 1.2-1.38, the present study is not insufficiently powered. However, in the presence of either significant genotypic or phenotypic heterogeneity, study power may be significantly diminished. As an example, it is plausible that certain SNPs may either be associated with only some disease subtypes or have opposite effects across JIA subtypes, as we have demonstrated with the GWS SNPs in our study. Thus, the method of hsGWAS which considers JIA subtype heterogeneity boosts the study power. Indeed, we have demonstrated that we have sufficient power to detect genetic loci conferring medium effect sizes and we have uncovered novel loci associated with JIA. However, small sample size is still a limitation of our study and we anticipate that with onging recruitment, we will have larger sample sizes including all JIA subtypes that will improve study power and will likely to yield more shared genetic loci among JIA subtypes.

## Methods

### Study Population

The Juvenile Idiopathic Arthritis (JIA) cohort was recruited in the USA, Australia, and Norway and comprised of a total of 1385 patients with onset of arthritis at < 16 years of age. JIA diagnosis and JIA subtype were determined according to the International League of Associations for Rheumatology (ILAR) revised criteria^59^ and confirmed using the JIA CalculatorTM software^51^ (http://www.jra-research.org/JIAcalc/), an algorithm-based tool adapted from the ILAR criteria. Prior to standard quality control (QC) procedures and exclusion of non-European ancestry, the JIA cohort was comprised of 464 case subjects from Texas Scottish Rite Hospital for Children (TSRHC; Dallas, Texas, USA) and The Children’s Mercy Hospitals and Clinics (CMHC; Kansas City, Missouri, USA) of self-reported European ancestry; 196 subjects from the Children’s Hospital of Philadelphia (CHOP; Philadelphia, Pennsylvania, USA); 221 subjects from the Murdoch Children’s Research Institute (MCRI; Royal Children’s Hospital, Melbourne, Australia); 504 subjects from Oslo University Hospital (OUH; Oslo, Norway). Age and gender matched control subjects were identified from the CHOP-CAG biobank and ascertained by exclusion of any patient with any ICD9 codes for disorders of autoimmunity or immunodeficiency (http://eicd9.com/).

### Ethics statement

Research Ethics Boards of CHOP and other collaborating centers approved this study, and written informed consent was obtained from all subjects (or their legal guardians).

### Genotyping

Genomic DNA was extracted from peripheral blood and sample quality control (QC) filtering before and after genotyping were performed using standard methods as described previously^60^. All samples in the cohort were genotyped at CAG on HumanHap550 and 610 BeadChip arrays (Illumina, CA). The SNPs genotype was called using BeadStudio (Illumina, CA) using the default parameters. To minimize population stratification, only individuals of self-reported European ancestry and further confirmed by principal-component analysis (PCA) were included for the present study. Details of the PCA are provided below.

### Sample and SNP QC

SNPs with a low genotyping rate (< 95%) or low MAF (< 0.01) or those significantly departing from the expected Hardy-Weinberg equilibrium (HWE; P < 1 × 10−^6^) were excluded. Samples with low overall genotyping call rates (< 95%) or determined to be of outliers of European ancestry by PCA (detailed below) were removed. In addition, one of each pair of related individuals as determined by identity-by-state analysis (PI_HAT > 0.1875) was excluded, with cases preferentially retained where possible.

### Principal-component analysis

To assess ethnicity, we merged our SNP data together with HapMap to do a principal-component analysis (PCA). We took the set of SNPs common to both datasets and pruned them using ‘plink --indep-pairwise 50 10 0.2’. We conducted PCA on the dataset with the pruned SNPs via PLINK^61^. K-means clustering was used to group subjects into distinct populations of origin and subjects of European ancestry were identified. Then PCA was similarly conducted among subjects of European ancestry in our dataset again to derive within population structure.

### Genome-wide SNP imputation

We used SHAPEIT^62^ for whole-chromosome pre-phasing and IMPUTE2 for imputation to the 1KGP-RP (https://mathgen.stats.ox.ac.uk/impute/impute_v2.html, June 2014 haplotype release). For both, we used parameters suggested by the developers of the software and described elsewhere^62-64^. Imputation was done for each 5-Mb regional chunk across the genome, and data were subsequently merged for association testing. Prior to imputation, all SNPs were filtered using the criteria described above. We filtered out poorly imputed SNP with INFO score < 0.8.

### Association analyses

We performed whole-genome association testing using post-imputation genotype probabilities with the software SNPTEST (v2.5)^65^. We used logistic regression to estimate odds ratios and betas, 95% confidence intervals and P values for trend, using additive coding for genotypes (0, 1 or 2 minor alleles). For autosomal regions, we used a score test, whereas for regions on ChrX we used the ChrX-specific SNPTEST method Newml. QC was performed directly after association testing, excluding any SNPs with an INFO score of < 0.80, HWE P < 1 × 10−^6^, and MAF < 0.01 (overall). In all analyses, we adjusted for both gender and ancestry by conditioning on gender and the first 9 principal components derived from PLINK PCA, which yielded a λ_GC_ values were within acceptable limits for all disease sub-type cohorts.

The extent of population structure was assessed by visual inspection of a quantile-quantile (QQ) plot of the test statistics and by calculating the inflation factor lambda. This was done for multiple version of the association analysis, in order to assess whether inflation has sufficiently decreased.

We performed stepwise conditional association analyses at each GWS locus. The genotype of the index SNP at each GWS locus was included as a covariate in the additive logistic regression model of the score test using SNPTEST. Any lead SNP with association P-value < 1e-06 was considered as an independent signal and was included as a covariate in the next round of conditional analyses. We performed such stepwise conditional association analyses until the P-value of the index SNP is greater than 1e-06.

### Heterogeneity-sensitive meta-analysis

To identify genetic loci that are associated with multiple sub-types of JIA and to determine the subtype-combination that each locus is most strongly associated, the “h.types” and “h.traits” methods implemented in the R statistical software package ASSET was applied to do an exhaustive disease-subtype model search, which has been described in detail in our and others’ publications^66,67^. Briefly, the different combinations of JIA subtypes were exhaustively enumerated and tested for association with each locus. The combination that yielded the best association statistics were selected as the best disease-subtype model. In our analyses, the “h.types” and “h.traits” methods yielded similar results. The DLM method was for multiple testing correction across all subtype combinations.

### Fine-mapping

Fine-mapping was performed using FINEMAP v1.3.1, a software for discerning causal SNPs of complex traits and calculating their effect and heritability contribution^68^. GWAS summary statistics and SNP Pearson’s correlation matrixes calculated from the genotyped data on the same individuals were used as input for FINEMAP. A shotgun stochastic search for “large *p*” regression was applied to attain a robust control. ^50^ For each GWS SNP, the summary statistics from GWAS of combination of JIA subtypes yielding the best association statistics were applied. Default parameter setting was adopted with the SNPs with the maximum number of allowed causal SNPs to be five. Likely causal SNPs with posterior probability greater than 0.2 and hsGWAS P-value < 1e-04 were selected.

### Pathway and PPI network analysis

The over-representation pathway analysis with 18 candidate genes at 16 GWS loci were conducted at web portal WEB-based gene set analysis Toolkit (http://www.webgestalt.org/)^69^. The PPI network visualization analysis and Competitive pathway enrichment analysis based on genome-wide summary-level data were performed with GSA-SNP2 (https://sites.google.com/view/gsasnp2) ^70^. Highly correlated adjoining genes were combined based on LD information of 1000G European population. The default setting of GSA-SNP2 was adopted to define each gene’s transcript region and its 20 kb upstream/downstream and its gene region. The collection of gene-set databases include BioCarta, KEGG, Reactome and PID by Molecular Signatures Database (MSigDB). The STRING database is the resource for the network construction. The significance threshold was defined as Q-value < 0.05 after multiple-testing correction. The significance of gene-score < 0.005 and Q-value < 0.05 was chosen for constructing global visual network.

### JIA subtype clustering

We performed unsupervised clustering on the JIA subtypes with the 16 GWS SNPs and 28 GMS SNPs. We generated the heatmap based on the hsGWAS P-value using R package Pheatmap. We also generated a t-SNE cluster plot according to the P-values and Betas of each SNP in the hsGWAS using the t-Distributed Stochastic Neighbor Embedding algorithm (t-SNE, Van Der Maaten et al. 2008), an unsupervised, non-linear technique primarily being used to produce clustering visualizations by reducing high-dimensional data. R package Rtsne was used.

### HLA imputation

The SNPs within the HLA region, spanning 29-34 Mb on chr6 of the human genome build hg19, were extracted after the SNP array data have been QCed. All JIA cases and controls were imputed together using software SNP2HLA http://www.broadinstitute.org/mpg/snp2hla/), with the T1DGC (the type 1 diabetes genetics consortium) reference panel. The frequency of HLA allele at a 2-digit level were compared between cases and controls for each JIA subtype, and the odds ratio and the association P-value were derived from the chi-square test of the 2×2 table.

## Data Availability

To have access to the data reported in this manuscript, please contact the corresponding author Dr. Hakon Hakonarson.

## Acknowledgements

We are grateful to all the patients for their participation in the study.

## Competing interests

The authors declare that the research was conducted in the absence of any commercial or financial relationships that could be a potential conflict of interest.

## Integrative Genetics Analysis of Juvenile Idiopathic Arthritis Identifies Novel Loci

## Notes

### Competing Interest Statement

The authors have declared no competing interest.

### Funding Statement

The study was supported by Institutional Development Funds from the Children's Hospital of Philadelphia to the Center for Applied Genomics, The Children's Hospital of Philadelphia Endowed Chair in Genomic Research to HH, and the grant U01-HG006830 (NHGRI-sponsored eMERGE Network) to HH.

